# Developing and Validating a New Web-Based Tapping Test for Measuring Distal Bradykinesia in Parkinson’s Disease

**DOI:** 10.1101/2020.06.30.20141572

**Authors:** Noreen Akram, Haoxuan Li, Aaron Ben-Joseph, Caroline Budu, David Gallagher, Jonathan P Bestwick, Alastair J Noyce, Cristina Simonet

**Author notes:** **Corresponding author:** Dr Cristina Simonet, Preventive Neurology Unit, Wolfson Institute of Preventive Medicine, Barts and the London School of Medicine and Dentistry, Queen Mary University of London, London, UK. These authors contributed equally to this work (Noreen Akram, Haoxuan Li). **Funding agencies:** There was no specific funding awarded for this study. **Financial Disclosures:** Dr. Noyce is funded by the Barts Charity. Dr. Noyce reports additional grants from Parkinson’s UK, Virginia Kieley benefaction, grants and non-financial support from GE Healthcare, and personal fees from Profile, Roche, Biogen, Bial and Britannia, outside the submitted work. Dr. Simonet is funded by Fundación Alfonso Martín Escudero, Spain. No other disclosures were reported.

## Abstract

**Background:** Disability in Parkinson’s disease (PD) is measured by standardised scales including the MDS-UPDRS, which are subject to high inter and intra-rater variability and fail to capture subtle motor impairment. The BRadykinesia Akinesia INcoordination (BRAIN) test is a previously validated keyboard tapping test, evaluating proximal upper-limb motor impairment. Here, a new Distal Bradykinesia Tapping (DBT) test was developed to assess distal upper-limb function. Kinetic parameters of the test include kinesia score (KS20, key taps over 20 seconds), akinesia time (AT20, mean dwell-time on each key) and incoordination score (IS20, variance of travelling time between key taps).

**Objective:** To develop and validate a new keyboard-tapping test to assess distal motor function in PD patients.

**Methods:** The DBT test was validated in 45 PD patients and 24 controls, alongside the BRAIN test. Test scores were compared between groups and correlated with MDS-UPDRS-III scores. 10 additional PD patients were recruited to assess the DBT test in monitoring motor fluctuations.

**Results:** All three parameters discriminated between patients and controls, with KS20 performing best, yielding 75% sensitivity for 85% specificity; area under the receiver operating characteristic curve (AUC) = 0.87. Combination of both the DBT and BRAIN tests improved discrimination (AUC=0.91). KS20 and AT20 correlated with MDS-UPDRS-III (Pearson’s r=-0.49, p<0.001 and r=0.54, p<0.001, respectively). The DBT test detected subtle changes in motor fluctuation states, which were not reflected clearly by MDS-UPDRS-III sub-scores.

**Conclusion:** The DBT test is a user-friendly method of assessing distal motor dysfunction in PD, possibly permitting longitudinal monitoring of PD motor complications.

## INTRODUCTION

Bradykinesia is an integral feature of Parkinson’s disease (PD) and relates to the ‘slowness of movement initiation with progressive reduction in speed and amplitude of repetitive actions’, as defined by the Queen Square Brain Bank Criteria (1). It is often assessed using the Movement Disorder Society Unified Parkinson’s Disease Rating Scale (MDS-UPDRS) part III (motor score) (2). Although the MDS-UPDRS-III is a comprehensive assessment, the integer scale prevents detection of subtle motor changes (3, 4) and is subject to high inter and intra-rater variability (5). Hence, there is a need for objective and consistent methods of assessing motor dysfunction.

The BRadykinesia Akinesia INcoordination (BRAIN) test is a previously validated tool for detecting upper-limb motor dysfunction (6, 7). This online tapping test requires participants to alternately tap the ‘S’ and ‘;’ keys on a computer keyboard using one index finger, as fast and accurately as possible, for 30 seconds (6). The test captures proximal motor impairment, as movement arises at the level of the elbow and shoulder. Existing literature suggests that proximal and distal movements are governed by two distinct neural pathways (8, 9). This possibly explains why, as a diagnostic test, the BRAIN test historically demonstrates a relatively low detection rate (sensitivity) for PD (58-65%) given high specificity (81-88%) (10). Additionally, the BRAIN test requires significant hand-eye coordination, which may be unsuitable for patients with visual impairment and/or severe tremor (10). To address these gaps, a new Distal Bradykinesia Tapping (DBT) test was developed.

## METHODS

### DBT test

The DBT test is an online tapping test, compatible with laptops and computers, accessed by participants using unique tokens (via https://predictpd.com/en/braintest).

Participants were instructed to repeatedly tap the down arrow key with their left index finger, as fast as possible for 20 seconds, whilst simultaneously depressing the left arrow key with their left middle finger. These instructions were then repeated for the right hand. These instructions stabilise the wrist and forearm, isolating movement to the index finger metacarpal joint, thereby giving a true measurement of distal bradykinesia.

Three kinetic parameters were generated by the DBT test: kinesia score (KS20), the number of keystrokes in 20 seconds reflecting speed; akinesia time (AT20), average dwell time that keys are depressed reflecting akinesia; and incoordination score (IS20), variance of travelling time between keystrokes reflecting rhythm.

### Participants

In the first stage of the study, PD patients fulfilling the Queen Square Brain Bank criteria were recruited from the Movement Disorder clinic at the Royal London Hospital between February and August 2019. They were frequency-matched with non-neurological controls. For the second stage, PD patients taking dopaminergic treatment and experiencing motor fluctuations were recruited from the same clinic.

Participants were seated in front of a computer/ laptop, where they independently undertook the DBT and BRAIN test. Total MDS-UPDRS-III scores were recorded for PD patients by trained individuals (NA &ABJ). The patients’ clinical state was recorded, with ‘On’ defined as a functional state when there is a good response to medication, and ‘Off’ defined as a poor functional state despite taking medication or after the symptomatic effect of medication had passed. Additionally, to investigate the presence of a learning effect, seven of the healthy controls completed the DBT test five times within a 3-hour period.

The second part of the study evaluated the use of the DBT test in assessing motor fluctuations, through home-visits. Patients performed the DBT and BRAIN test, alongside MDS-UPDRS-III assessment, carried out by the same trained neurologist (CS). Assessments coincided with patients’ motor fluctuations. Four patients were invited to complete the DBT test on further occasions at home, according to their subjective impressions about being ‘On’ or ‘Off’, for longer monitoring of fluctuations.

### Statistical analysis

Normality was assessed using D’Agostino and Pearson test/ Shapiro-Wilk test. Descriptive statistics were calculated for all three parameters (KS20, AT20, IS20), with mean being reported for normally distributed data and median for not normally distributed data. DBT test scores in patients and controls were compared using the unpaired t-test and Mann-Whitney U test for parametric and non-parametric data respectively. Receiver operating characteristic (ROC) curves were generated using Wilson/Brown method, determining sensitivity and specificity of parameters. Logistic regression and ROC curves defined AUC values for combination analysis of DBT and BRAIN test variables. Relationship between test parameters and MDS-UPDRS-III were assessed using Pearson’s correlation and Spearman’s rank correlation. In controls, one-way repeated measures ANOVA was used to detect a learning effect. Paired t-tests and Wilcoxon matched-pairs signed rank tests investigated whether the DBT test and MDS-UPDRS-III could differentiate between fluctuation states. The significance level for all calculations was set as p<0.0025 (derived by Bonferroni calculation to reduce type 1 error). All data were analysed using GraphPad Prism version 8.0.2 and IBM SPSS Version 26.

All participants were appropriately consented. Ethics approval was granted by the Queen Square Research Ethics Committee (09/H0716/48).

## RESULTS

Forty-five PD patients and twenty-four frequency-matched controls were included in the main analysis of the first part of the study. Eight PD patients were excluded due to significant tremor and comorbidities, such as rheumatoid arthritis. There was no significant difference in age, sex and ethnicity between PD subjects and controls. *Table 1* summarises the demographic data for participants. In the second stage, ten additional PD patients were recruited for monitoring motor complications (mean age in years ± SD: 61.90 ± 7.25, mean disease duration with PD in years ± SD: 9.1 ± 5.22 and gender distribution: 50% male and 50% female). One patient was excluded due to cognitive impairment. One patient only contributed towards UPDRS-III assessment of fluctuations due to insufficient left hand DBT data, and another patient was excluded from UPDRS-III analyses due to only the ‘On’ state being captured during the home visit.

**Table 1.** Baseline characteristics of PD patients and controls.

|  | PD | Controls |
| --- | --- | --- |
| **Number** | 45 | 24 |
| **Mean Age (SD)** | 67.4 (8.8) | 66.0 (11.8) |
| **Gender** |  |  |
| - Female | 19 (42%) | 16 (67 %) |
| - Male | 26 (58%) | 8 (33 %) |
| **Ethnicity** |  |  |
| - White British | 19 (42%) | 11 (46%) |
| - South Asian | 16 (36%) | 6 (25%) |
| - Black British | 4 (9%) | 2 (8%) |
| - Other | 6 (13%) | 5 (21%) |
| **Mean Yrs since diagnosis (SD)** | 6.3 (4.9) | - |
| **Education** |  |  |
| - Primary | 3 (7%) | - |
| - Secondary | 18 (40%) | - |
| - Higher | 8 (18%) | - |
| - Further | 16 (35%) | - |
| **Occupation** |  |  |
| - Professional | 21 (47%) | - |
| - Non-professional skilled | 6 (13%) | - |
| - Non-professional/non-skilled | 18 (40%) | - |
| **Hand dominance\*** |  |  |
| - Right | 20 (95%) | 21 (87%) |
| - Left | 1 (5%) | 3 (13%) |
| **Most affected side** |  |  |
| - Right | 15 (33%) | - |
| - Left | 28 (62%) | - |
| -Equally | 2 (4%) | - |
| **Levodopa** |  |  |
| - Yes | 42 (93%) | - |
| - No | 3 (7%) | - |
| **Mean minutes since levodopa dose (SD)** | 157.8 (109.2) | - |
| **On/Off\*\*** |  |  |
| - On | 40 (95%) | - |
| - Off | 2 (5%) | - |

\* Handedness data was only available for 21 PD participants
\*\* On/off refers to the question in the MDS-UPDRS, which asks whether patients taking levodopa could notice the effect of medication at the time of examination

Associations between DBT test parameters with age, gender and handedness was assessed in control subjects (*see Table 2*). Neither age nor gender overtly affected the test parameters. Handedness had an effect on KS20, AT20 and IS20 in controls only.

**Table 2.** Analysis of characteristics that influence KS20, AT20 and IS20 in controls.

|  |  | Mean<br>KS20* | p-Value | Mean<br>AT20* | p-value | Median<br>IS20* | p-value |
| --- | --- | --- | --- | --- | --- | --- | --- |
| **Mean Age** | 66.0 | r=-0.27 | 0.20<sup>a</sup> | r=0.16 | 0.46<sup>a</sup> | r=-0.12 | 0.59<sup>b</sup> |
| **Gender** |  |  |  |  |  |  |  |
| - Female | 16 | 80.6 | 0.87<sup>c</sup> | 132.3 | 0.40<sup>c</sup> | 1376 | 0.65<sup>d</sup> |
| - Male | 8 | 81.4 |  | 117.5 |  | 1271 |  |
| **Handedness** |  |  |  |  |  |  |  |
| - Dominant | 24 | 84.3 | <0.001<sup>e</sup> | 114.0 | <0.001<sup>e</sup> | 783.7 | <0.001<sup>f</sup> |
| - Nondominant | 24 | 77.5 |  | 140.8 |  | 1108 |  |

KS20, kinesia score; AT20, akinesia time; IS20, incoordination score \*Mean and medians given except for associations with age where correlation coefficient (r) is given. <sup>a</sup>Pearson;
<sup>b</sup>Spearman; <sup>c</sup>Unpaired t-test; <sup>d</sup>Mann-Whitney, <sup>e</sup>Paired t-test, <sup>f</sup>Wilcoxon test

### Validating the DBT test in patients and controls

All three parameters discriminated between patients and controls. KS20 was the best discriminator, with 75% sensitivity for 85% specificity and an AUC of 0.87 (*see Table 3 and Figure 1*). The corresponding sensitivities for 85% specificity for AT20 and IS20 were 54% and 46%, with respective AUC’s of 0.81 and 0.79 *(see Table 3 and Figure 1)*. The combination of DBT parameters improved discrimination with an AUC of 0.88, and the combination of both DBT and BRAIN test parameters gave an AUC of 0.91. A moderate correlation was found for KS20 and AT20 against total MDS-UPDRS-III scores (Pearson’s r=-0.49, p<0.001 and r=0.54, p<0.001 respectively) (*see Figure 2*). Repeat testing in seven of the controls did not reveal any learning effect in KS20 (p=0.53), AT20 (p=0.58) or IS20 (p=0.24), using one-way repeated measures ANOVA.

**Table 3.**
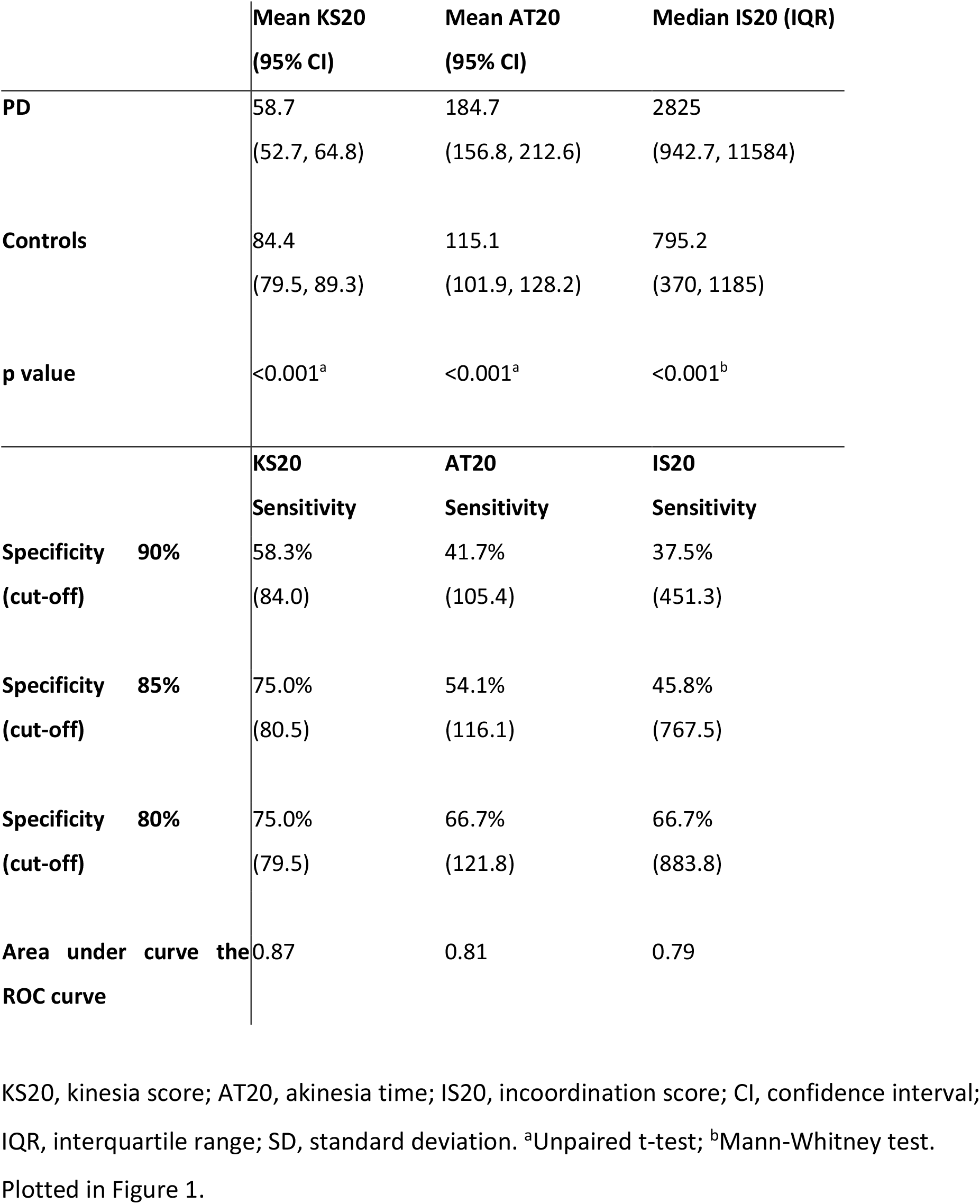
Comparison of KS20, AT20, and IS20 between patients and controls and corresponding ROC analysis.

**Figure 1.**
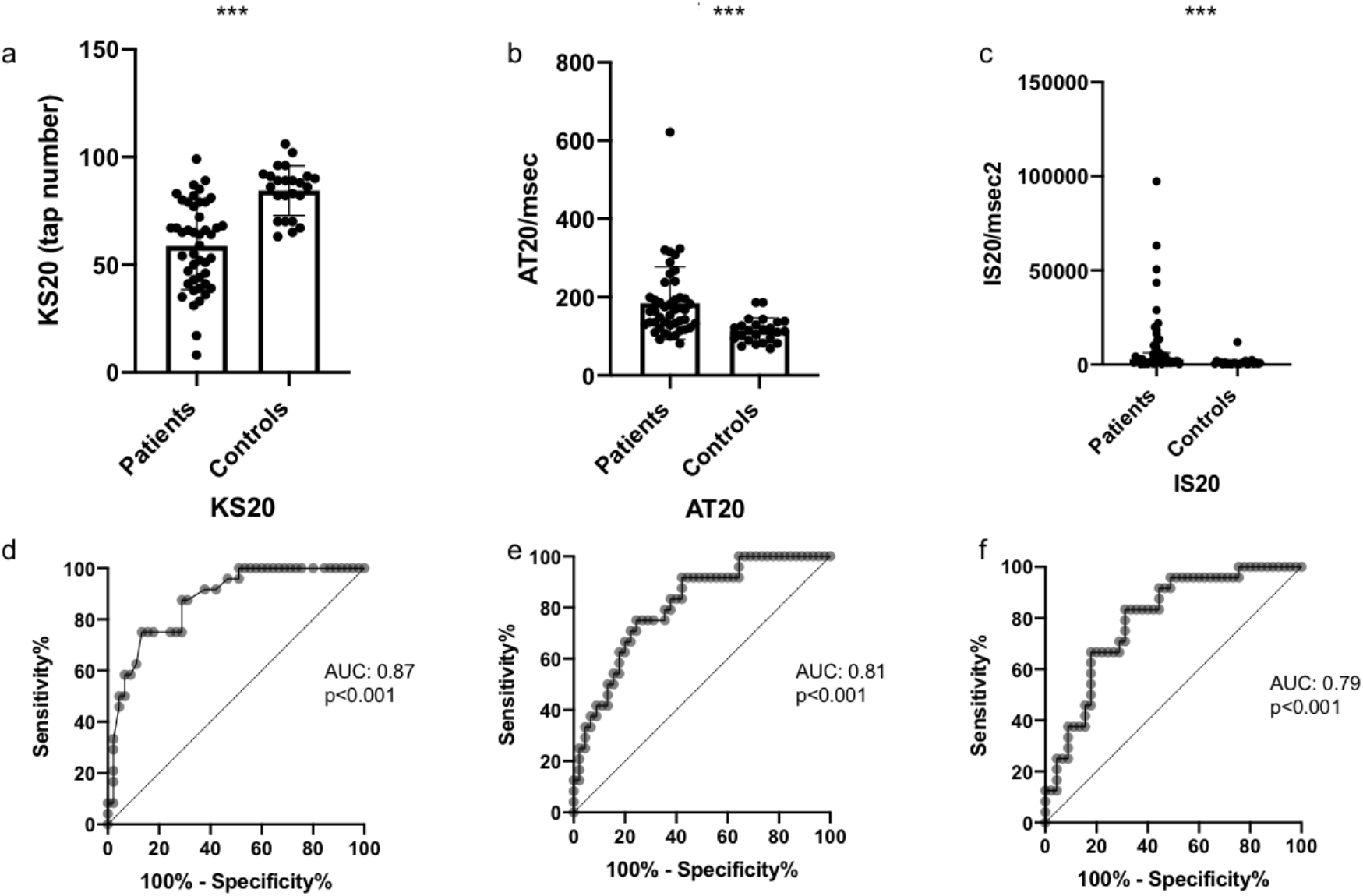
Comparison of KS20, AT20 and IS20 in PD patients and controls. Spread of a) KS20, b) AT20 (mean and SD) and c) IS20 (median and IQR) for patients and controls. Receiver operating curves for d) KS20, e) AT20 and f) IS20. Plotted in Figure 1.

**Figure 2.**
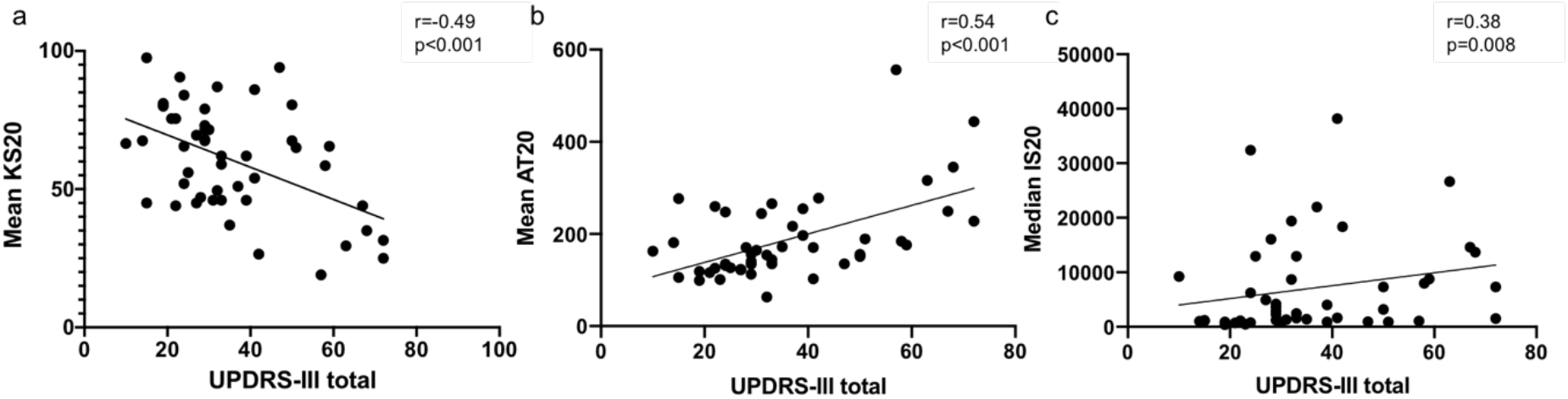
Correlation of KS20, AT20 and IS20 with total MDS-UPDRS-III. a) Moderate negative correlation with KS20 and UPDRS. b) Moderate positive correlation seen with AT20 and UPDRS. c) no significant correlation seen with IS20 and UPDRS.

### Monitoring motor fluctuations

The DBT test provided suggestive evidence for a difference between patients’ ‘On’ and ‘Off’ states using KS20 and IS20 (p=0.05 and p=0.04, respectively; s*ee Table 4*). Contrastingly, the finger tapping sub-score of the MDS-UPDRS-III showed less evidence of being able to differentiate between fluctuation states (p=0.10). Results for four patients who completed the tapping tests more than twice, found KS20 to be the most consistent parameter (*see Figure 3*). In **patient 1**, KS and AT scores from the DBT and BRAIN test demonstrated ‘on-off’ fluctuations. In **patient 2**, the DBT test demonstrated a progressive decrease in ‘On’ state KS20 scores throughout the day, whilst ‘Off’ state KS20 scores remained relatively constant, potentially reflecting a diurnal variation in PD symptoms. This pattern was not demonstrable using the BRAIN test. Of note, in **patient 4**, the KS20 score did not rise following the third LD dose, possibly reflecting an additional ‘no on’ or ‘delayed on’, which was not detected using the BRAIN test.

**Table 4.** Comparison of KS20, AT20 and IS20 between patients’ ‘On’ and ‘Off’ states.

| **Parameter** | **PD 'Off'** | **PD 'On'** | **p-value** |
| --- | --- | --- | --- |
| **Mean KS20 in taps (95% CI)** | 62.78<br>(50.06, 75.50) | 71.78<br>(61.49, 82.07) | 0.05<sup>a</sup> |
| **Mean AT20 in msec (95% CI)** | 155.0<br>(118.3, 191.7) | 153.1<br>(120.6, 185.7) | 0.88<sup>a</sup> |
| **Median IS20 in msec<sup>2</sup> (IQR)** | 3452<br>(1833, 20178) | 1232<br>(845.3, 10017) | 0.04<sup>b</sup> |

KS20, kinesia score; AT20, akinesia time; IS20, incoordination score; CI, confidence interval; IQR, interquartile range. <sup>a</sup>Two-tailed paired t-test, <sup>b</sup>Wilcoxon matched-pairs signed rank test.

**Figure 3:**
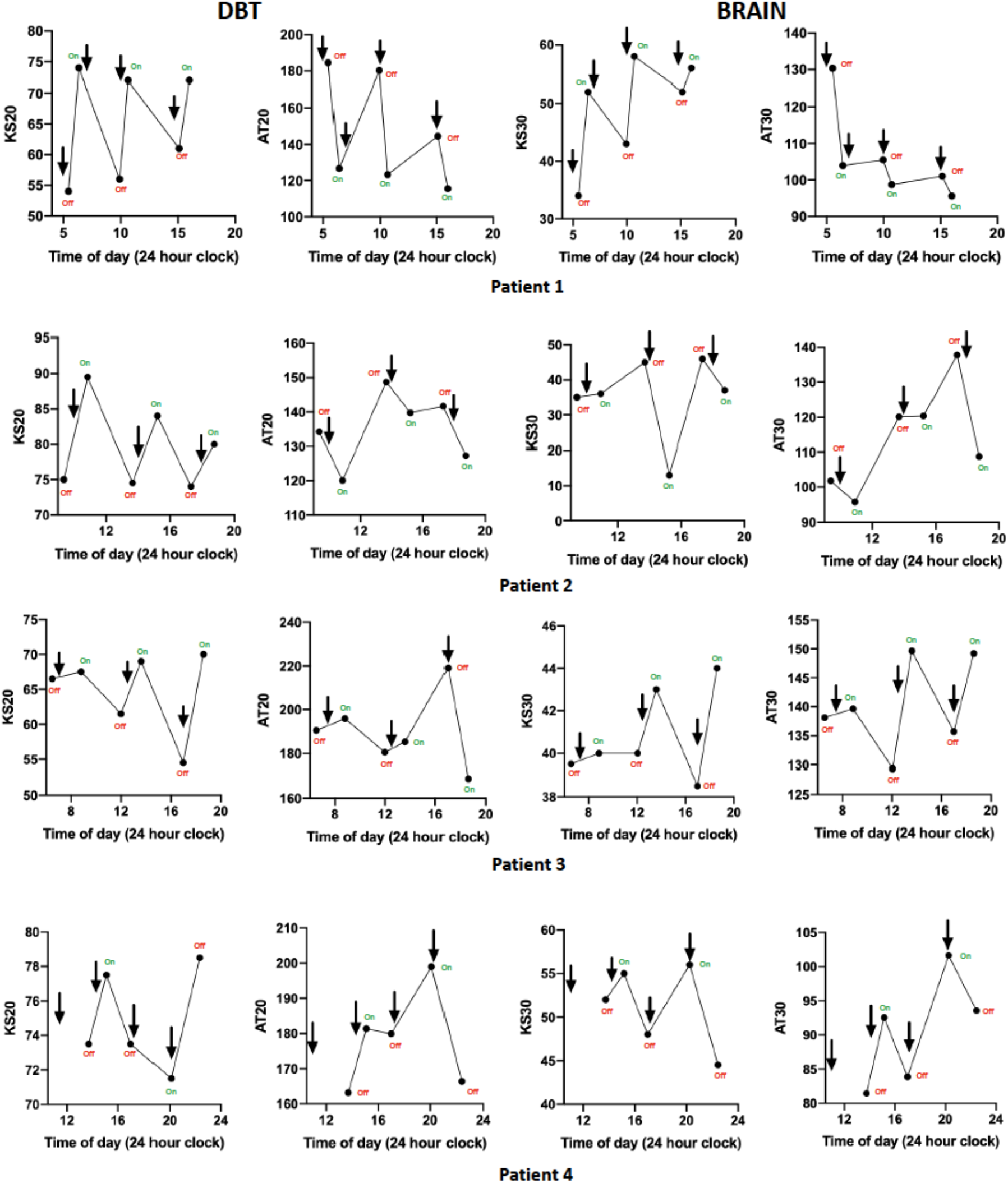
Repeat testing in 4 PD patients with predictable motor fluctuations using the DBT and BRAIN test. Dots represent when the test was completed, and arrows denote the time when levodopa was taken. KS20 (DBT test) and KS30 (BRAIN test) scores are expected to increase in the ‘On’ state, whereas AT20 and AT30 scores are expected to decrease in the ‘On’ state.

## DISCUSSION

All three DBT test parameters distinguished patients from controls, with KS20 being the best discriminator. Both KS20 and AT20 demonstrated good sensitivity levels for high specificity levels, thereby reducing false-positive rates. The combination of all three DBT test parameters improved diagnostic performance, and the integration of both DBT and BRAIN test considerably improved AUC to 0.91. These findings support our original hypothesis about the tests assessing proximal and distal bradykinesia respectively. KS20 and AT20 from the DBT test correlated with the MDS-UPDRS-III, and may therefore be useful surrogate markers for assessing disease severity. The DBT and BRAIN test may be utilised in tandem, improving diagnostic ability and providing complementary information on proximal and distal bradykinesia, possibly guiding PD management.

Age or gender did not influence the results. Participants from varied backgrounds were able to conduct the test, thereby increasing applicability. Lastly, learning effect was not strongly demonstrable for any DBT parameters, thus supporting its use for longitudinal monitoring.

The DBT test (KS20/ IS20) appear able to detect subtle changes in ‘On’ and ‘Off’ states, whilst the finger tapping sub-score of the MDS-UPDRS-III was less able to identify such discrepancies. Rating scales can be a crude measure of motor function and thus may not detect subtle symptom oscillations as demonstrated in the fluctuation graphs produced by the DBT test. These findings however remain exploratory and warrant further investigation in a larger sample size. Overall, the DBT and BRAIN test demonstrated distinct sensitivities in detecting subtle motor fluctuations for individual patients, reinforcing the idea that distal and proximal movements may be differentially affected in PD (11-14). It is evident that both tapping tests are useful in objectively capturing daily symptom oscillations, and providing a clearer understanding of patients’ subjective interpretations of fluctuation states.

One limitation of the DBT test is the lack of amplitude measurement, a key feature of bradykinesia. Moreover, these tests measure speed and rhythm of upper limb movement and does not address rigidity and tremor. Likewise, this test may not be possible in context of cognitive impairment, marked tremor or arthropathy, due to difficulties in completing the task.

The DBT test is a simple cost-effective method, devoid of specialised equipment. Moreover, the test uniquely captures isolated distal bradykinesia, allowing refinement of therapeutic options. The DBT test addresses limitations faced by the BRAIN test, eliminating complex hand-eye coordination and interference of ‘paradoxical kinesia’ (actively finding the ‘s’ and ‘;’ keys could have a visual-clue-role and transiently improve bradykinesia) (15).

Future directions would be to study the ability of both tests to detect early motor manifestations in population-based longitudinal studies, such as PREDICT-PD study (16).

## CONCLUSION

The novel DBT test offers a reliable and objective method of capturing true distal bradykinesia. It is a simple and user-friendly tool which can be employed in both clinical and home settings and serves as a supplementary clinical tool for remote monitoring of PD motor complications.

## Data Availability

The authors confirm that the data supporting the findings of this study are available within the article [and/or] its supplementary materials.

## AUTHOR’S ROLES

1) Research project: A. Conception, B. Organisation, C. Execution. 2) Statistical Analysis: A. Design, B. Execution, C. Review and Critique. 3) Manuscript Preperation: A. Writing of the first draft, B. Review and Critique.

NA: 1A, 1B, 1C. 2A, 2B, 3A

HL: 1A, 1B, 1C. 2A, 2B, 3A

ABJ: 2C, 3B

CB: 2C, 3B DG: 2C, 3B JPB: 2C, 3B

AJN: 1B, 2C, 3B

CS: 1A, 1B, 2A, 2C, 3B

## ETHICAL COMPLIANCE STATEMENT

We confirm that we have read the Journal’s position on issues involved in ethical publication and affirm that this work is consistent with those guidelines. The authors confirm that all participants gave verbal and written consent for this work. Ethics approval was granted by the Queen Square Research Ethics Committee (09/H0716/48).

## REFERENCES

1. Hughes AJ, Daniel SE, Kilford L, Lees AJ. Accuracy of clinical diagnosis of idiopathic Parkinson’s disease: a clinico-pathological study of 100 cases. J Neurol Neurosurg Psychiatry. 1992;55(3):181–4.

2. Goetz CG, Tilley BC, Shaftman SR, Stebbins GT, Fahn S, Martinez-Martin P, et al. Movement Disorder Society-sponsored revision of the Unified Parkinson’s Disease Rating Scale (MDS-UPDRS): scale presentation and clinimetric testing results. Mov Disord. 2008;23(15):2129–70.

3. Taylor Tavares AL, Jefferis GS, Koop M, Hill BC, Hastie T, Heit G, et al. Quantitative measurements of alternating finger tapping in Parkinson’s disease correlate with UPDRS motor disability and reveal the improvement in fine motor control from medication and deep brain stimulation. Mov Disord. 2005;20(10):1286–98.

4. Regnault A, Boroojerdi B, Meunier J, Bani M, Morel T, Cano S. Does the MDS-UPDRS provide the precision to assess progression in early Parkinson’s disease? Learnings from the Parkinson’s progression marker initiative cohort. J Neurol. 2019;266(8):1927–36.

5. Hasan H, Athauda DS, Foltynie T, Noyce AJ. Technologies Assessing Limb Bradykinesia in Parkinson’s Disease. J Parkinsons Dis. 2017;7(1):65–77.

6. Noyce AJ, Nagy A, Acharya S, Hadavi S, Bestwick JP, Fearnley J, et al. Bradykinesia-akinesia incoordination test: validating an online keyboard test of upper limb function. PLoS One. 2014;9(4):e96260.

7. Shribman S, Hasan H, Hadavi S, Giovannoni G, Noyce AJ. The BRAIN test: a keyboard- tapping test to assess disability and clinical features of multiple sclerosis. J Neurol. 2018;265(2):285–90.

8. Takahashi K, Best MD, Huh N, Brown KA, Tobaa AA, Hatsopoulos NG. Encoding of Both Reaching and Grasping Kinematics in Dorsal and Ventral Premotor Cortices. J Neurosci. 2017;37(7):1733–46.

9. Fattori P, Breveglieri R, Raos V, Bosco A, Galletti C. Vision for action in the macaque medial posterior parietal cortex. J Neurosci. 2012;32(9):3221–34.

10. Hasan H, Burrows M, Athauda DS, Hellman B, James B, Warner T, et al. The BRadykinesia Akinesia INcoordination (BRAIN) tap test: capturing the sequence effect. 2018.

11. Weiss PH, Dafotakis M, Metten L, Noth J. Distal and proximal prehension is differentially affected by Parkinson’s disease. The effect of conscious and subconscious load cues. J Neurol. 2009;256(3):450–6.

12. Dafotakis M, Fink GR, Allert N, Nowak DA. The impact of subthalamic deep brain stimulation on bradykinesia of proximal and distal upper limb muscles in Parkinson’s disease. J Neurol. 2008;255(3):429–37.

13. Fellows SJ, Noth J. Grip force abnormalities in de novo Parkinson’s disease. Mov Disord. 2004;19(5):560–5.

14. Delong MR, Georgopoulos AP, Crutcher MD, Mitchell SJ, Richardson RT, Alexander GE. Functional organization of the basal ganglia: contributions of single-cell recording studies. Ciba Found Symp. 1984;107:64–82.

15. Schlesinger I, Erikh I, Yarnitsky D. Paradoxical kinesia at war. Mov Disord. 2007;22(16):2394–7.

16. Noyce AJ, Bestwick JP, Silveira-Moriyama L, Hawkes CH, Knowles CH, Hardy J, et al. PREDICT-PD: identifying risk of Parkinson’s disease in the community: methods and baseline results. J Neurol Neurosurg Psychiatry. 2014;85(1):31–7.

